# Nucleobindin-2 (NUCB2) gene transcription activity and nesfatin-1 levels in correlation with anthropometric and biochemical parameters in type 2 diabetes mellitus patient groups in Vietnam

**DOI:** 10.1101/2021.09.27.21263999

**Authors:** Duc Minh Nguyen, Minh Thi Nguyen, Mao Van Can, Huong Ngoc Thu Trinh, Linh Bao Ngo, Thuy Thi Bich Vo, Minh Ngoc Nghiem

## Abstract

**Introduction:** Nucleobindin-2 (NUCB2) was identified as a DNA/Ca2+ binding protein with multiple functions in humans. Prohormone convertase-mediated NUCB2 processing produced nesfatin-1 - a biologically active. Nesfatin-1, an 82-amino acid peptide, was extracted from the N-terminus of nucleobindin-2. Recently, it was described as an anorexia peptide related to weight loss, malnutrition, and appetite regulation in type 2 diabetes mellitus patients.

**Research design and methods:** In this study, we collected samples and divided them into groups of patients with long-term type 2 diabetes and newly diagnosed type 2 diabetes group. Serum nesfatin-1 level and mRNA NUCB2 gene expression level of the groups were analyzed and compared with those of the healthy group.Biometric parameters and biochemical indices were also analyzed to determine the correlation with nesfatin-1 level.

**Results:** Levels of nesfatin-1 were found to be higher in the newly diagnosed group than in the other groups. Similar results were also reported in the analysis of mRNA NUCB2 gene expression by Realtime-PCR. Meanwhile, no significant difference was found in both analyzes of nesfatin-1 levels and NUCB2 mRNA expression in subjects with long-term type 2 diabetes compared with the control group. This result can be explained by the effects of long-term treatment. In the correlation of anthropometric parameters and biochemical indices, nesfatin-1 exhibited a significant correlation with BMI (r=0.569), HbA1c (r=-0.468), HDL-C (r=0.731), LDL-C (r=-0.482), Creatinine serum (r=0.525), and Creatinine urine (r=0.592), with p<0.001, in regression analysis.

**Conclusions:** These results indicate that the serum nesfatin-1 level and the NUCB2 mRNA gene expression level may be associated with newly diagnosed type 2 diabetes in Vietnamese patients. However, more specific studies with larger sample sizes were still needed in future studies.

## Introduction

Diabetes was recognized as the “pandemic” of the 21st century affecting millions of people worldwide. According to statistics of the International Diabetes Federation (IDF 2018), the world has about 425 million people with diabetes, of which over 90% have type 2 diabetes and this trend is increasing [1]. Type 2 diabetes mellitus (T2DM) was a long-term metabolic disorder characterized by high blood sugar (glucose) levels, insulin resistance, and relative insulin deficiency [2]. Common symptoms include extreme thirst, frequent urination, and unexplained weight loss, frequent feelings of hunger, fatigue, muscle weakness, and slow healing of wounds or bruises [3]. Type 2 diabetes was mainly caused by obesity and lack of exercise [4] and some people are more genetically at risk than others [2]. In the past 10 years, the rate of increase of diabetes in Vietnam was 211%, 3 times higher than that of the world (70%). And Vietnam also was among the 10 countries with the highest rate of increase in diabetes patients in the world with the patient rate increasing by 5.5% per year [5, 6]. According to estimates, Vietnam currently has 3.53 million people “living” with diabetes and at least 80 people die from related complications every day. The number of people infected with the disease in Vietnam will be reached to be 6.3 million by 2045.

Nucleobindin (NUCB2) was found in the plasma membrane and neuroplasma. Some prohirmone convertase enzymes of NUCB2 such as PC3/1 and PC2 converts NUCB2 to nesfatin-1 (1-82 aa), nesfatin-2 (85-163 aa) and nesfatin-3 (166-396aa) [14, 16, 19]. NUCB2 has a characteristic constitution of functional domains, such as a signal peptide, a Leu/Ile rich region, two Ca^2+^ binding EF-hand domains separated by an acidic amino acid-rich region, and a leucine zipper [22, 23]. In humans, the NUCB2 gene length was 55 kb with 14 exons and 13 introns. The translation region of the NUCB2 gene was described in exon-3. Nesfatin-1 was translated in region between exon-3 and 5 of NUCB2 gene [16, 17]. However, recent studies have shown that NUCB2 was also expressed in other peripheral tissues as the stomach, pancreas, reproductive organs, and adipose tissues [14, 20, 21].

Nesfatin-1 was discovered by Oh-I for the first time in 2006, it showed that nesfatin-1 was secreted by peripheral tissues, central and peripheral nervous systems (7). Nesfatin-1 length polypeptides have 82 amino acid which derived from the precursor protein NUCB2. The structure of nesfatin-1 has three parts: N-terminal (N23), Middle part (M30), and C-terminal (C29). The middle part was determined as an active part of it. It was important for the physiological effects of nesfatin-1 in the anorexic effect [16,17,18]. In the study of Gonzalez R (2009), the homology of nesfatin-1 was up to 85% between humans and other mammal species [12]. The play role of nesfatin-1 was as an appetite suppressant and was jointed in the balanced of energy homeostasis related with food and water intake without leptin gene (8). Specifically, nesfatin-1 enzyme was selected by peripheral adipose tissue, gastric mucosa, pancreatic endocrine beta cells, and testis tissue. It can pass through the blood-brain barrier after secretion [9,10,11,12,13,14].

In previous studies, nesfatin-1 has been suggested to be involved in type 2 diabetes mellitus (T2DM) pathogenesis by stimulating free acid utilization. A significant decrease in fasting plasma nesfatin-1 levels in T2DM and polycystic ovary syndrome (PCOS) patients has also been confirmed. This could be caused by impaired insulin sensitivity and suggests that nesfatin-1 might be inhibited by insulin resistance, hyperglycemia, and hyperinsulinemia [15]. The plasma level of nesfatin-1 was not increased by acute stress [24, 25]. However, the serum nesfatin-1 levels have been found to be higher in patients with depression than in controls [26].

In Vietnam, follow to our knowledge, no studies have been conducted on diabetes patients about nesfatin-1 levels and the expression of the NUCB2 gene. It is very important to understand whether nesfatin-1 can be used as a supportive tool in the diagnosis and treatment of diabetes in Vietnam patients. Thus, in this study, we conducted to measure nesfatin-1 concentration levels and mRNA expression of NUCB2 levels in adipose tissues by Realtime-PCR. The correlation between the relative expression of NUCB2, nesfatin-1 levels, and some clinicopathological parameters to evaluate its clinical significance.

## Materials and Methods

### Patients

A cross-sectional study was conducted on a total of 65 type 2 diabetic mellitus patients and 30 control subjects (17 male and 13 female, age 54.83 ± 9.56 yr). Those two groups of diabetic patients were divided by the duration of diabetes, type 2 diabetes mellitus (T2DM) group was 37 patients (15 male and 22 female, age 62.95± 7.09 yr) with over 4 years, newly diagnosed type 2 diabetes mellitus (nT2DM) group was included 28 patients (11 male and 17 female, age 60.54 ± 7.98 yr) with under 3 years. All patients were randomly selected from those who visited the Endocrinology Department of 19-8 Hospital, Ministry of Public Security, Vietnam, between January and December 2020. The diagnose of type 2 diabetes mellitus was made according to the American Diabetes Association criteria which were based on the criteria using a fasting glucose level ≥7.0 mmol/L. Clinical exclusion criteria included type 1 diabetes mellitus, history of endocrine disorders, pregnancy, surgery or trauma in recent times, heart failure, and cancer. All the people visited 19-8 hospital for regular physical examination were selected as the controls who had no family history of diabetes or other endocrine disorders. All people in this study were Vietnamese and consented to join. The present study was conducted based on the principles of the Declaration of Helsinki and approved by the Ethics Committee of Vietnam Military Medical University and 19-8 Hospital, Ministry of Public Security, Vietnam.

### Anthropometry

Anthropometric measurement was performed in the morning, before breakfast. Bodyweight and height were calculated by a scale and a wall-mounted stadiometer (0.5kg and 0.5cm, respectively). Body Mass Index (BMI) was a person’s weight in kilograms divided by the square of height in meters. Waist and hip circumference were calculated by standard method. The waist-to-hip ratio was calculated as waist measurement divided by hip measurement (W/H).

### Biochemical parameters

After an overnight fast, blood samples were collected and immediately centrifuged and were frozen at −80 C until assayed. Plasma glucose levels were measured with the glucose-oxidase technique and glycosylated hemoglobin (HbA1c) with anion-exchange HPLC. Levels of plasma total cholesterol, Creatinin Serum, Creatinine Urine, high-density lipoprotein cholesterol (HDL-C), and low-density lipoprotein cholesterol (LDL-C) were drawn in lithium-heparin vacuum tubes and analyzed enzymatically using an autoanalyzer.

### Serum nesfatin-1 assay

The serum nesfatin-1 levels were detected by using a commercial enzyme liked immunosorbent assay kit from Biomatik, Wilmington, US with the linear range: 31.25 pg/mL-5000 pg/mL.

### Adipose biopsy

Subcutaneous abdominal periumbilical adipose tissue biopsy samples were obtained under sterile conditions and local anesthesia with 1% Lidocaine. A biopsy needle with a small incision was used to obtain tissue samples. Samples were treated with liquid nitrogen to flash-freeze the tissue in 1 minute. The flash-frozen adipose tissue was cryopulverized 2 times, stored at −80°C until extraction and analysis.

### Analysis of mRNA NUCB2 expression by Realtime - PCR

Total RNA was isolated from adipose tissue (approximately 85-100 mg) using Trizol Total RNA was isolated from adipose tissue (90 mg) by Trizol reagent (Invitrogen, USA) follow to the manufacturer’s guide. Treat with DNase I (Invitrogen, USA), synthesis first-strand cDNA from purified RNA by SuperScript III (Invitrogen, USA). Quantitative Realtime - PCR was run with LC-Fast Start DNA SYBR Green I chemistry on a LightCycler 2.0 software (Roche, Switzerland). The expressions gene were analyzed by the comparative Ct method in relation to the GAPDH levels. The sequences for NUCB2 were: sense, 5’-GCCAGAACGTGTTACGAGTC-3’ and antisense, 5’-GTCCTCCACCTCATGTTCAG-3’. The relative amounts of the mRNA were quantified using the second derivative maximum method of the light-cycler software.

### Statistical analysis

Statistical analysis were analyzed with R and R-studio software [27, 28]. Data were expressed as mean ± SD or frequency (%). One-way ANOVA with post hoc (least significant difference) analysis to assess for differences in body composition, anthropometric, metabolic, and hormonal parameters among the T2DM patients, nT2DM patients, and controls group. The correlation of serum nesfatin-1 with other clinical characteristics (included: BMI, Creatinine Serum, Creatinine Urine, HDL-C, LDL-C, Total Cholesterol, HbA1c, and Fasting Blood Glucose) was determined using Pearson correlation analysis, *p* < 0.05 was considered significant.

## Results

### The clinical characteristics of the patient and control subjects

The clinical characteristics and laboratory findings of all subgroups were shown in Table 1. The highest range of age was given in the T2DM group (62.95 ± 7.09). And this level was determined to decrease gradually when compared with the nT2DM and the Control group (60.54 ± 7.98 and 54.83 ± 9.56, respectively). While both the Body mass index (BMI) and the Waist hip ratio (WHR) were significantly higher in patients groups (T2DM: 23.25 ± 2.05; 0.94 ± 0.05 and nT2DM: 22.61 ± 3.42; 0.93 ± 0.06) compared with the control group (22.07 ± 3.01 and 0.86 ± 0.03).

**Table 1.**
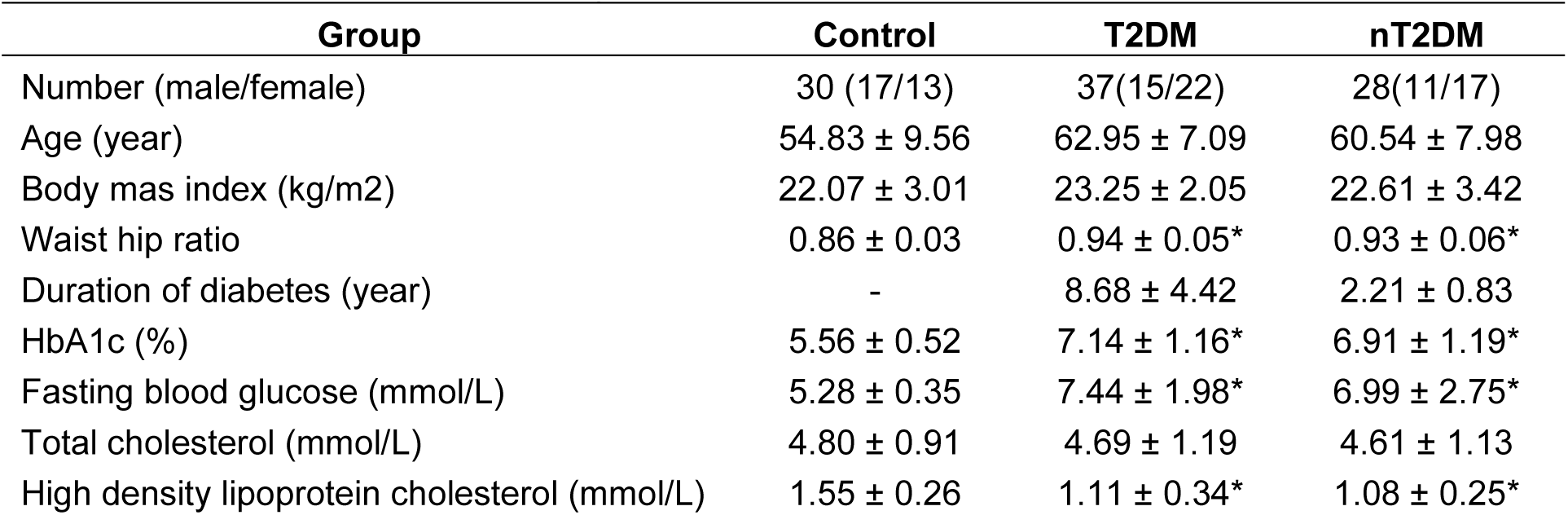
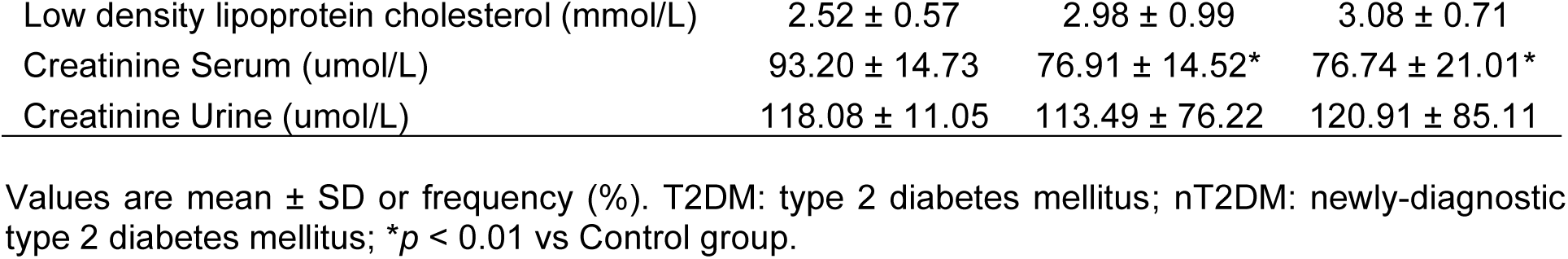
Demographic, anthropometric, and metabolic characteristics of diabetic mellitus patients and control subjects.

The duration of diabetes in the T2DM group approximately was 8 years, with a minimum range was 4 years and a maximum was 20 years (8.68 ± 4.42). In nT2DM patients, the 3 years diabetes time of this group was accounted for half of the total patients, included: 7 patients in 1 year, 8 patients in 2 years, and 13 patients in 3 years, its mean 2.21 ± 0.83.

In the frequency of glycosylated hemoglobin (HbA1c) unit, T2DM patients also were at the highest level (7.14 ± 1.16) compared to Control (5.56 ± 0.52) and nT2DM (6.91 ± 1.19). The fasting blood glucose was significantly higher in the T2DM group than in the nT2DM and control groups (7.44 ± 1.98 versus 6.99 ± 2.75 and 5.28 ± 0.35 mmol/L, *p* < 0.01). Which was an important index to identify the presence of diabetes (FBG ≥ 7 mmol/L).

The index about the amount of cholesterol in the body was determined based on the LDL cholesterol, HDL cholesterol, and total cholesterol levels. Total cholesterol levels were significantly increased in the Control groups when compared with the T2DM and nT2DM (4.80 ± 0.91 vs 4.69 ± 1.19 and 4.61 ± 1.13 mmol/L), but there were no significant differences between groups. All three study groups achieved an ideal concentration of less than 200mg/dL (equivalent to >5.1 mmol/L), which showed no increase in blood cholesterol and a reduced risk of Coronary Artery Disease (CAD).

In HDL-C and LDL-C levels, although in both the T2DM and nT2DM groups, they have not yet decreased to a bad level or need to be cautious (HDL-C < 1.0 mmol/L and LDL-C > 3.3 mmol/L), however, these indexes are being evaluated as inferior to control group (HDL-C: 1.11 ± 0.34 and 1.08 ± 0.25 vs 1.55 ± 0.26 mmol/L; LDL-C: 2.98 ± 0.99 and 3.08 ± 0.71 vs 2.52 ± 0.57 mmol/L). Especially with the nT2DM group, if lack of early treatment, which can easily lead to worse conditions.

The test assessed indices of kidney function such as Creatinine Serum and Creatinine Urine was also analyzed, however, not have great influence among the 3 study groups.

### Serum nesfatin-1 concentration in patients with type 2 diabetes mellitus and newly type 2 diabetes mellitus

As shown in Figure 1, the level of serum nefatin-1 was significantly higher in the nT2DM group than T2DM and control group (*p*<0.05 vs Control). In T2DM patients, this value was slightly lower than people of control, however, no significant differences among them (*p* = 0.068).

**Figure 1.**
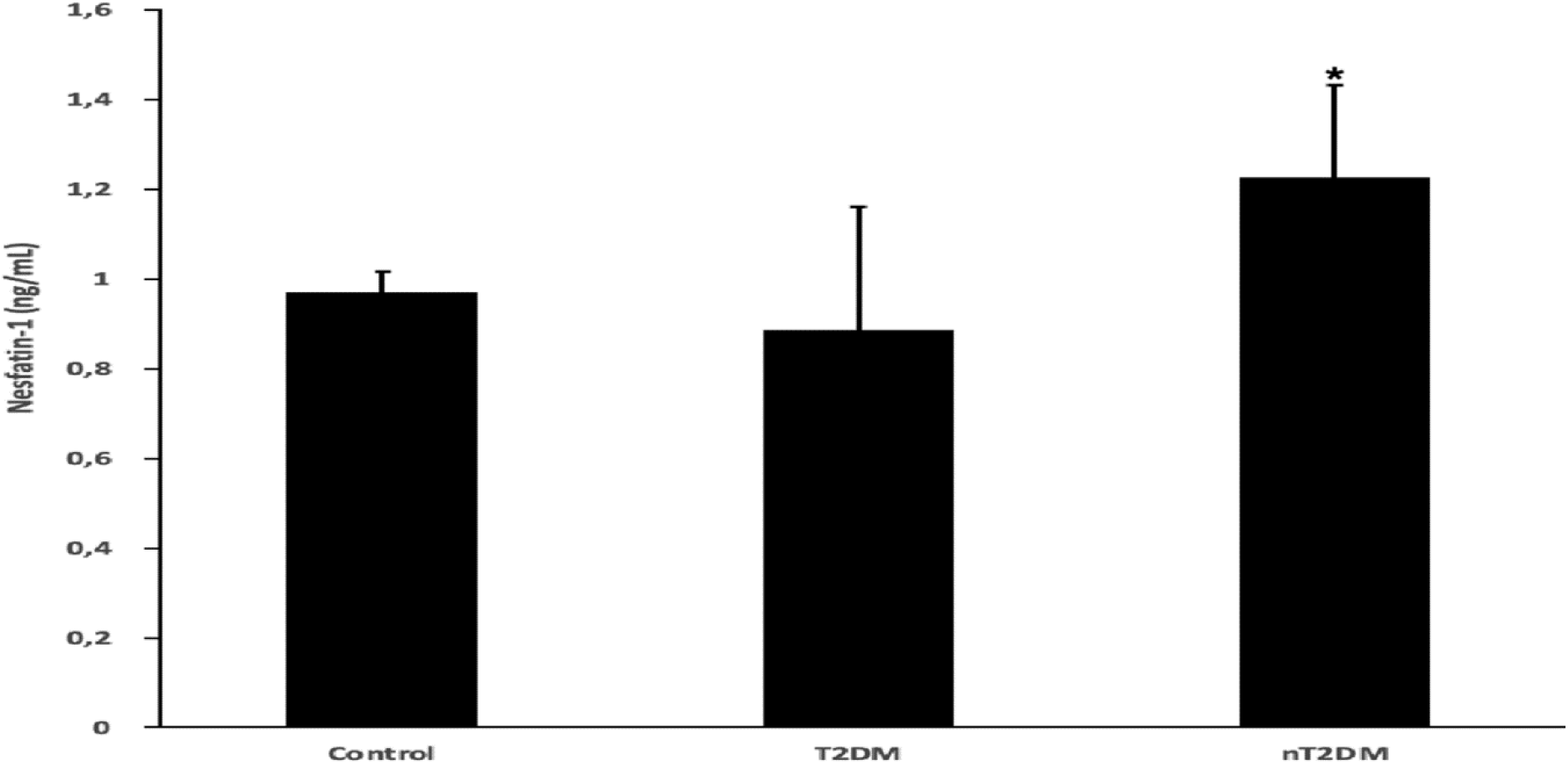
Concentration of nesfatin-1 (mean ± SEM) in 3 groups: Control (n=30) and patients with T2DM (n=37) and nT2DM (n=28); vs. Control group \**p* < 0.05.

### The correlation of serum nesfatin-1 with other clinical characteristics

The linear regression analysis was used to evaluate the correlation of serum nesfatin-1 and other metabolic parameters related to diabetes. The result showed that both of T2DM and nT2DM group, nesfatin-1 was positively correlated with BMI, WHR, Duration of diabetes, Total Cholesterol, HCL-C, Creatinin Serum, and Cretinin Urine. However, only correlation with duration of diabetes and total cholesterol were no significant differences in both groups (*p* = 0.102 and 0.764, respectively).

With other parameters such as HbA1c, FBG, and LDL-C, all confirmed to be negatively correlated with nesfatin-1, *p*-value < 0.05 for HbA1c and *p*<0.001 for LDL-C, no significant differences in the case of FBG (*p*=0.216 and 0.688) (Table 2).

**Table 2.**
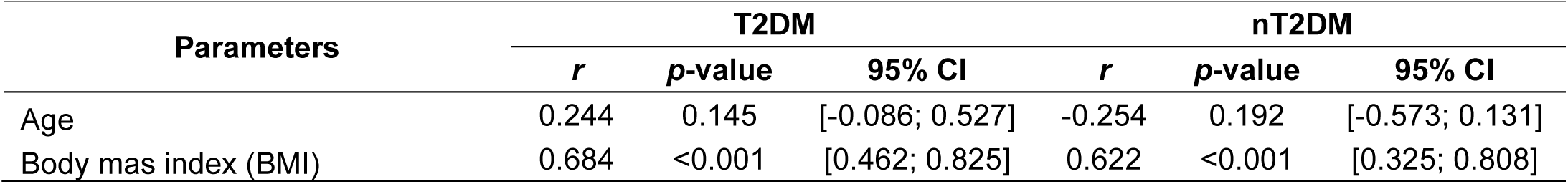
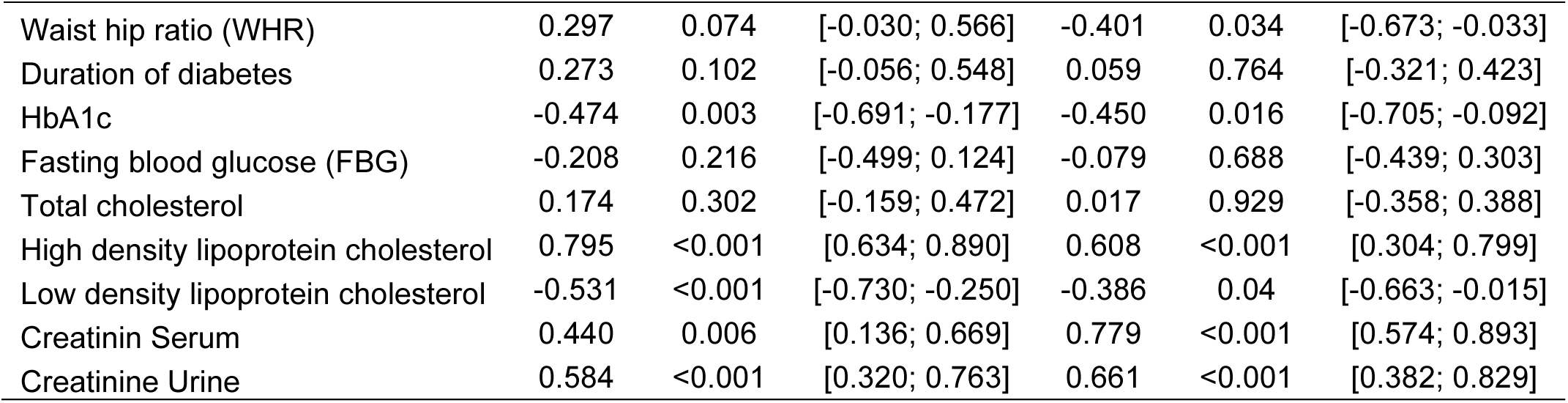
The correlation analysis results between clinical variables and serum nesfatin-1 levels in diabetes patient groups.

Based on the correlation between nesfatin-1 indexes and the parameters which were determined to have a significant difference with p<0.05 or <0.001, we conducted a multiple stepwise regression analysis with 6 units, such as HDL-C, LDL-C, Creatinine Serum, Creatinine Urine, BMI, and HbA1c with all of 65 patients in T2DM and nT2DM groups. The results are shown in Figure 2.

**Figure 2.**
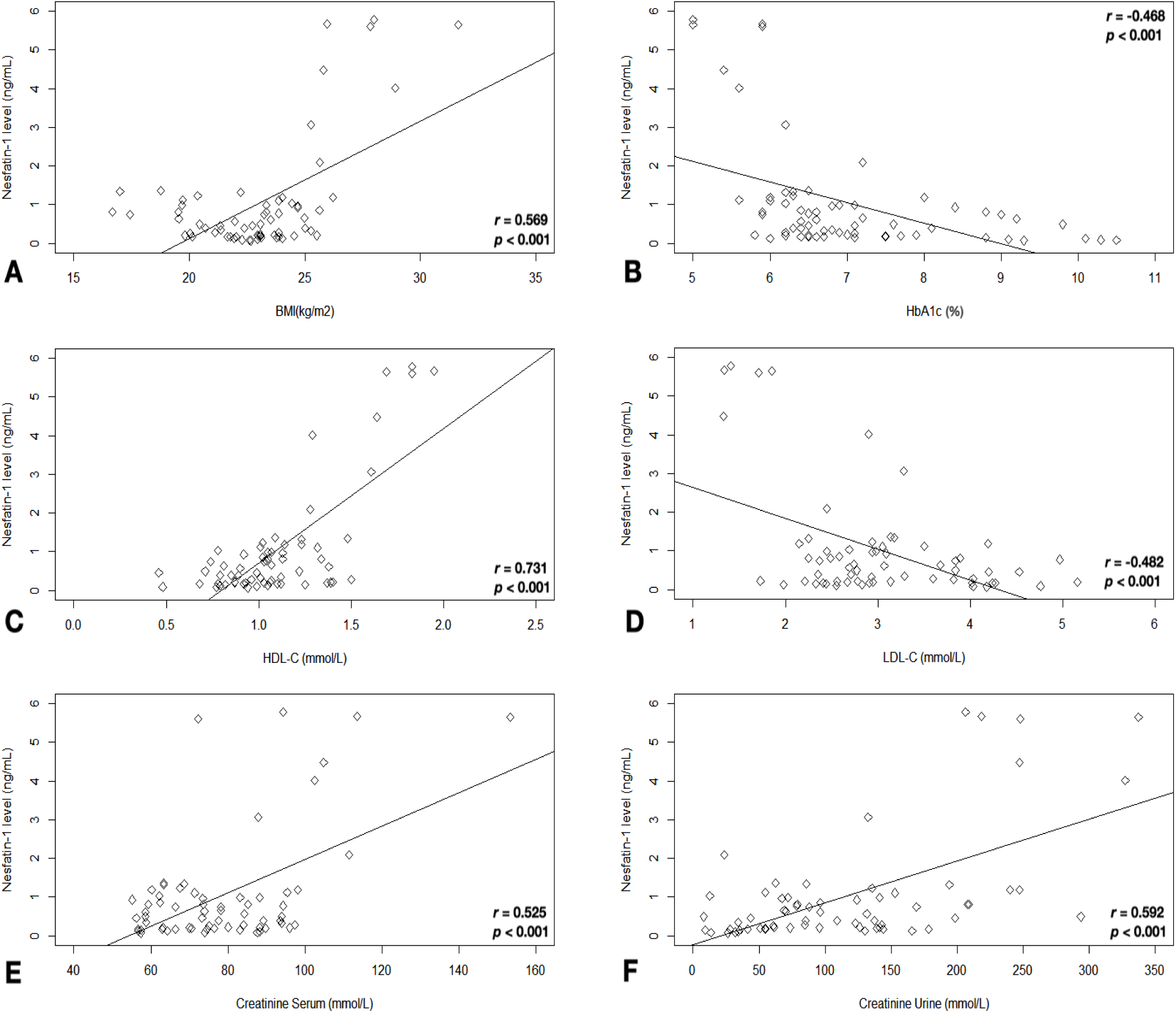
Correlations between nesfatin-1 with BMI (A), HbA1c (B), HDL-C (C), LDL-C (D), Creatinine serum (E) and Creatinine Urine (F) in diabetes patients.

### NUCB2 mRNA expression in adipose tissues

In this study, only a certain number of people agree to join the subcutaneous adipose tissue experiment (included: 5 people in the control group, 10 patients in the T2DM group, and 10 patients in the nT2DM group). Follow the result in Figure 3, NUCB2 mRNA expressions were lower in adipose tissues from nT2DM patients than those from controls and T2DM (by ∼1.2-fold and ∼1.5-fold respectively, *p*<0.01 vs Control). The expression of genes was consistent with nesfatin-1 levels in serum which was also markedly increased in nT2DM patients compared with controls and T2DM.

**Figure 3.**
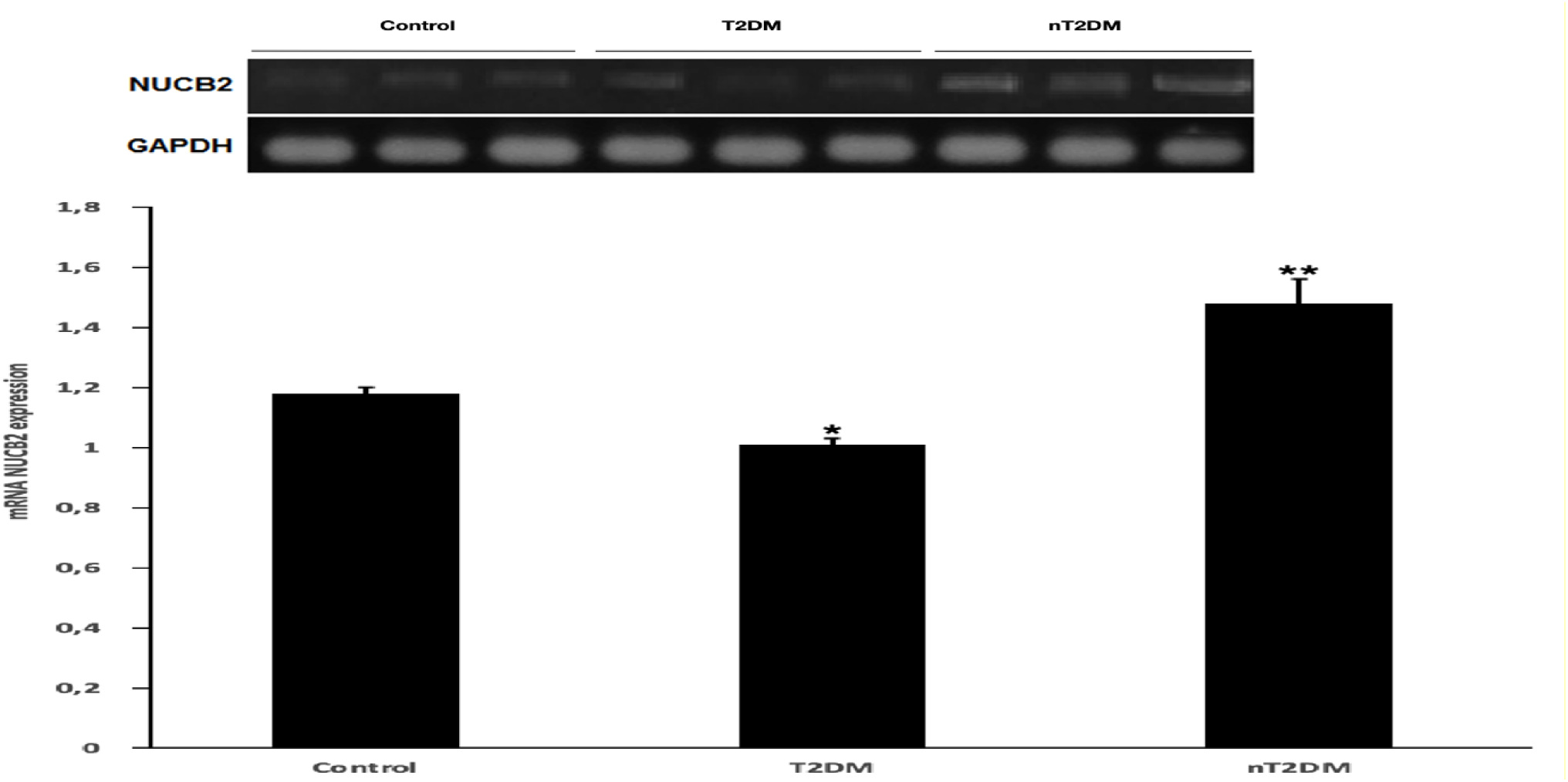
Real-time RT-PCR analysis of NUCB2 mRNA levels in human adipose tissues collected from Control subgroup (n=5) with T2DM (n=10) and nT2DM (n=10) subgroup. Data are means±SE; \**p*<0.05 \*\**p*<0.01 vs Control.

## Discussion

NUCB2/nesfatin-1 was first identified in several regions of the hypothalamus, and subsequently in peripheral tissues including adipocytes, gastric mucosa, pituitary gland, heart, and medulla oblongata of humans and rats (13). Recent studies have reported that nesfatin-1 was a potential candidate to treat type 2 diabetes mellitus with hypoglycemic effects in conditions of impaired glucose metabolism (29). It may also act in the brain to regulate insulin sensitivity (30) and increase insulin release in beta cells to respond with hyperglycemia (31).

In our study, the level of nesfatin-1 in the newly diagnosed type 2 diabetes group was higher than in the healthy group. This result was determined to be consistent with previous studies such as the study of Zhang, 2012 and Guo, 2013. This could be explained as a physiological response or a compensatory mechanism for the impaired insulin action of newly diagnosed type 2 diabetes patients (32 - 34).

However, in the study by Li, the fasting nesfatin-1 concentration (after 24 h of fasting) of type 2 diabetics was significantly lower than that of the healthy group (15). This result has also been shown to be similar in later studies (35 - 38). It was different with our data when the nesfatin-1 level of the subjects with long disease duration (8-20 years) was not significantly different from the control group. To account for these different results, there may be variations in the selection of patients in the experimental design, as well as in the experimental conditions or duration of the test time. For example, in our experiment, the majority of patients participating in the study were sampled in the morning, after overnight fasting (12 hours). Therefore, the interval after the last meal may not be enough to see a difference in nesfatin-1 levels between the 2 study groups, the long-term diabetic patients and the control group.

Besides, to further explain the results of our study, the nesfatin-1 level of the long-term type 2 diabetes group may be affected by medicines diabetes treatments. With the goal of reducing blood sugar, increasing insulin sensitivity, and controlling food intake, there are currently many classes of drugs commonly used in the treatment of diabetes, such as Metformin, Sulfonylureas, Meglitinides, Thiazolidinediones, DPP-4 inhibitors, GLP-1 receptor agonists, SGLT2 inhibitors or Insulin. Thus, the effects of these drugs on each patient’s concentration of nesfatin-1 were unavoidable.

In order to better clarify the correlation between serum nesfatin-1 level and some related parameters, we conducted analyzes by simple regression method on study subjects. With 3 anthropometric parameters studied (included: Age, BMI, and WHR), only the BMI parameter showed a significant difference in the positive correlation with nesfatin-1 level (p<0.001). Although there was no significant difference in the correlation between nesfatin-1 and age, however, our result was consistent with that reported in the study by Li (15). There is a further explanation of the results of the study reported on the Vietnamese group of patients, possibly because the habit of periodic health check-ups has not been established in young people, so most of the patients only conduct the health check-ups after retirement age (standard supported by the government). Therefore, the patient group identified with the type 2 diabetes disease in the study was relatively old, leading to the result that the correlation for the age group in our study is not clear. In addition, the parameters of BMI and WHRare also different from previous studies, which may be the result of differences between groups of people around the world.

In addition, the positive correlations and significant differences were reported in the two creatinine test indices. This result is equivalent to the study of Li (15). The reported negative correlation coefficient between nes-1 level with HbA1c, HDL-C and LDL-C is similar to some previous studies (15, 35). The above results suggest that these factors may contribute to the uniformity observed in the overall analysis of nesfatin-1 serum concentrations of type 2 diabetes patients.

To further clarify whether the NUCB2 gene expression level affects the serum nes-1 level in Vietnamese patients, we analyzed the mRNA NUCB2 gene expression level by Realtime-PCR. Previous studies have shown the of mRNA NUCB2 expression in peripheral tissues including heart, spinal cord, pancreas, stomach, muscle and adipose tissue (12, 13). They have important physiological roles in body weight and contribute to the pathophysiology of insulin resistance and related metabolic problems in obesity and diabetes. Guo Y found that mRNA NUCB2 and protein levels of muscle and adipose tissue were significantly increased in T2DM patients (32). Nesfatin-1 has been described to be able to cross the blood-brain barrier across an unsaturated membrane (10, 11). The expression of mRNA NUCB2 was established in the gastric mucosa higher 10 times more than in the brain, suggesting that the stomach was the major base of circulating nesfatin-1 (39). The expression of NUCB2 and nesfatin-1 was also released in adipocytes. This expression was mainly evident in subcutaneous adipose tissue cells (16).

In this study, the mRNA NUCB2 gene expression in Vietnamese patients with type 2 diabetes, reports have shown high expression levels in newly diagnosed type 2 diabetes group. While the expression level of NUCB2 gene transcription activity in the long-term disease group was observed to be slightly decreased compared with the control group. This may be explained by the fact that the long-term disease patients have been treated with different classes of drugs, which compensate for the lack of insulin. Thus, the mRNA NUCB2 expression level in this group was not different compared the heathly group.

Our study is limited by many factors, such as experimental design, participants, time and research funding, therefore, the sample size was relatively small, which leads to the inability to clarify the clear relationship between the data reported.

In conclusion, the novelty of our study was the first study to determine the nesfatin-1 concentration and the mRNA NUCB2 gene expression level in a group of Vietnamese patients with type 2 diabetes mellitus. It has similar with previous studies in the world. Moreover, the relationship between nesfatin--1 in serum with some anthropometric and biochemical parameters of the long-term disease and newly diagnosed diseases of Vietnamese patients was also published for the first time. The in-depth studies in a larger sample population is required. However, our findings also suggest some potential of using nesfatin-1 as an effective treatment for type 2 diabetes in Vietnamese patients in the future.

## Data Availability

None of the authors has any potential conflict of interests associated with this research.

## Acknowledgments

This work was supported by grants from the Graduate University of Sciences and Technology (GUST.STS.DDT2020-SH02), and the Vietnam Academy of Science and Technology (NCVCC40.03/21-21).

## References

1. Federation, Internation Diabetes. “IDF diabetes atlas ninth.” Dunia: IDF (2019).

2. “Causes of Diabetes”. National Institute of Diabetes and Digestive and Kidney Diseases. June 2014. Archived from the original on 2 February 2016. Retrieved 10 February 2016.

3. “Diagnosis of Diabetes and Prediabetes”. National Institute of Diabetes and Digestive and Kidney Diseases. June 2014. Archived from the original on 6 March 2016. Retrieved 10 February 2016.

4. “Diabetes Fact sheet N°312”. World Health Organization. August 2011. Archived from the original on 26 August 2013. Retrieved 2012-01-09.

5. Nguyen Hai Thuy, Tran Huu Dang. “Diabetes Update Document” (2013): 115–357.

6. Tran Huu Dang, Tran Thua Nguyen. “Diabetes Recommendations: Epidemiology, classification, and diagnosis”, Journal of the Vietnam Diabetes Endocrine Society (2016): 5–12.

7. Oh, Shinsuke, et al. “Identification of nesfatin-1 as a satiety molecule in the hypothalamus.” Nature 443.7112 (2006): 709–709.

8. Ayada, C. E. Y. L. A. N., Ü. M. R. A. N. Toru, and Y. A. S. E. M. İ. N. Korkut. “Nesfatin-1 and its effects on different systems.” Hippokratia 19.1 (2015): 4.

9. Stengel, Andreas, and Yvette Taché. “Nesfatin-1—role as possible new potent regulator of food intake.” Regulatory peptides 163.1-3 (2010): 18–23.

10. Pan, Weihong, Hung Hsuchou, and Abba J. Kastin. “Nesfatin-1 crosses the blood–brain barrier without saturation.” peptides 28.11 (2007): 2223–2228.

11. Price, Tulin O., et al. “Permeability of the blood–brain barrier to a novel satiety molecule nesfatin-1.” Peptides 28.12 (2007): 2372–2381.

12. Gonzalez, Ronald, Akansha Tiwari, and Suraj Unniappan. “Pancreatic beta cells colocalize insulin and pronesfatin immunoreactivity in rodents.” Biochemical and biophysical research communications 381.4 (2009): 643–648.

13. Stengel, Andreas, et al. “Identification and characterization of nesfatin-1 immunoreactivity in endocrine cell types of the rat gastric oxyntic mucosa.” Endocrinology 150.1 (2009): 232–238.

14. Garcia-Galiano, David, et al. “Expanding roles of NUCB2/nesfatin-1 in neuroendocrine regulation.” Journal of molecular endocrinology 45.5 (2010): 281–290.

15. Li, Qing-Chun, et al. “Fasting plasma levels of nesfatin-1 in patients with type 1 and type 2 diabetes mellitus and the nutrient-related fluctuation of nesfatin-1 level in normal humans.” Regulatory peptides 159.1-3 (2010): 72–77.

16. Pałasz, Artur, et al. “Nesfatin-1, a unique regulatory neuropeptide of the brain.” Neuropeptides 46.3 (2012): 105–112.

17. Yamada, Masanobu, et al. “Troglitazone, a ligand of peroxisome proliferator-activated receptor-γ, stabilizes NUCB2 (nesfatin) mRNA by activating the ERK1/2 pathway: isolation and characterization of the human NUCB2 gene.” Endocrinology 151.6 (2010): 2494–2503.

18. Aydin, Suleyman. “Role of NUCB2/nesfatin-1 as a possible biomarker.” Current pharmaceutical design 19.39 (2013): 6986–6992.

19. Williams, Paul, et al. “Expression of nucleobindin 1 (NUCB1) in pancreatic islets and other endocrine tissues.” Cell and tissue research 358.2 (2014): 331–342.

20. Kalnina, Zane, et al. “Molecular characterisation and expression analysis of SEREX-defined antigen NUCB2 in gastric epithelium, gastritis and gastric cancer.” European journal of histochemistry: EJH 53.1 (2009).

21. Suzuki, Shiho, et al. “Nucleobindin 2 in human breast carcinoma as a potent prognostic factor.” Cancer science 103.1 (2012): 136–143.

22. Miura, Keiji, et al. “Molecular cloning of nucleobindin, a novel DNA-binding protein that contains both a signal peptide and a leucine zipper structure.” Biochemical and biophysical research communications 187.1 (1992): 375–380.

23. Barnikol-Watanabe, Shitsu, et al. “Human protein NEFA, a novel DNA binding/EF-hand/leucine zipper protein. Molecular cloning and sequence analysis of the cDNA, isolation and characterization of the protein.” (1994): 497–512.

24. Goebel, Miriam, et al. “Restraint stress activates nesfatin-1-immunoreactive brain nuclei in rats.” Brain research 1300 (2009): 114–124.

25. Yoshida, Natsu, et al. “Stressor-responsive central nesfatin-1 activates corticotropin-releasing hormone, noradrenaline and serotonin neurons and evokes hypothalamic-pituitary-adrenal axis.” Aging (Albany NY) 2.11 (2010): 775.

26. Ari, Mustafa, et al. “High plasma nesfatin-1 level in patients with major depressive disorder.” Progress in neuro-psychopharmacology and biological psychiatry 35.2 (2011): 497–500.

27. The R Project for Statistical Computing. Available at: https://www.r-project.org/. Accessed: 1 Aug 2019.

28. Lê, Sébastien, Julie Josse, and François Husson. “FactoMineR: an R package for multivariate analysis.” Journal of statistical software 25.1 (2008): 1–18.

29. Su, Yijing, et al. “The novel function of nesfatin-1: anti-hyperglycemia.” Biochemical and biophysical research communications 391.1 (2010): 1039–1042.

30. Yang, Mengliu, et al. “Nesfatin-1 action in the brain increases insulin sensitivity through Akt/AMPK/TORC2 pathway in diet-induced insulin resistance.” Diabetes 61.8 (2012): 1959–1968.

31. Nakata, Masanori, et al. “Nesfatin-1 enhances glucose-induced insulin secretion by promoting Ca2+ influx through L-type channels in mouse islet β-cells.” Endocrine journal (2011): 1102080532-1102080532.

32. Guo, Y., et al. “Increased nucleobindin-2 (NUCB2) transcriptional activity links the regulation of insulin sensitivity in Type 2 diabetes mellitus.” Journal of endocrinological investigation 36.10 (2013): 883–888.

33. Zhang, Zhihong, et al. “Increased plasma levels of nesfatin-1 in patients with newly diagnosed type 2 diabetes mellitus.” Experimental and clinical endocrinology & diabetes 120.02 (2012): 91–95.

34. Khalili, Soodabeh, et al. “NUCB2/Nesfatin-1: a potent meal regulatory hormone and its role in diabetes.” Egyptian Journal of Medical Human Genetics 18.2 (2017): 105–109.

35. Algul, S., Y. Ozkan, and O. Ozcelik. “Serum nesfatin-1 levels in patients with different glucose tolerance levels.” Physiological research 65.6 (2016).

36. Dai, Rongfeng, et al. “Relation of serum and vitreous nesfatin-1 concentrations with diabetic retinopathy.” Journal of clinical laboratory analysis 31.5 (2017): e22105.

37. Liu, Fupeng, et al. “Decreased plasma nesfatin-1 level is related to the thyroid dysfunction in patients with type 2 diabetes mellitus.” Journal of Diabetes Research 2014 (2014).

38. Tang, Jiazhen, et al. “Changes in the serum levels of nesfatin-1 in type 2 diabetic patients with nephropathy: 407.” Diabetes/Metabolism: Research & Reviews 31 (2015).

39. Li, Ziru, et al. “Peripheral effects of nesfatin-1 on glucose homeostasis.” PloS one 8.8 (2013): e71513.

